# Historic Redlining and Hypertensive Disorders of Pregnancy: A Retrospective Cohort Study

**DOI:** 10.1101/2025.06.16.25328662

**Authors:** Patricia J Goedecke, Rachel Nelson, Hanah Walker, Anna Belle Gallaher, Kendra Hotz, Angela Nakahara, Lauren Camp, Charisse Madlock, Saunak Sen, Irma Singarella, Giancarlo Mari, Claire Simpson, Audris Mockus

**Affiliations:** University of Tennessee Health Science Center; Rhodes College; University of Iowa; University of Memphis; University Hospitals; University of Tennessee Knoxville

## Abstract

**Objectives:** The discriminatory practice of redlining, coding certain neighborhoods to receive less access to homeowner loans, has been suggested to be strongly associated with poor health outcomes. This study aims to: (1) investigate the relationship between historic redlining and hypertensive disorders of pregnancy (HDP); (2) identify current social determinants of health (SDOH) associated with historic redlining; (3) explore the potential network of relationships among these SDOH and HDP.

**Study Design:** Retrospective cohort study comparing pregnancies of women residing in historically redlined neighborhoods (grade D) with women in neighborhoods graded A-C who delivered in Memphis, Tennessee, from 2019-2020. The primary outcome was the development of HDP, measured using risk ratios and multivariable logistic modeling. We used prior literature to identify current SDOH associated with redlining. Where these were continuous, we modeled univariate linear relationships with neighborhood grades. In a third, exploratory analysis, we modeled relationships among current SDOH related to redlining and HDP applying Bayesian network analysis (BNA).

**Findings:** HDP prevalence differed significantly across historic redlining grade levels, with univariate risk ratios of (.59, .55, and .77) for grades A through C, respectively, with grade D as baseline (p = 0.0129). Multivariable logistic modeling showed a 22% increased risk (CI 1.00, 1.50) of HDP for each neighborhood grade. The literature identified eight current SDOHs potentially associated with neighborhood grades; of these, seven were continuous. Univariate regression found that 5 of the 7 had linear associations with neighborhood grades. Our network analysis did not find an association between HDP and current SDOH using BNA. It did show population-level chronic hypertension to be associated with scarcity of current residential loans (2011-2020) and property abandonment.

**Discussion:** Our findings support the growing body of evidence suggesting lasting effects of disinvestment in historically redlined neighborhoods.

**Disclosures:** Audris Mockus, Saunak Sen, Irma Singarella, and Giancarlo Mari provided guidance on the development of study goals and methods. Patricia Goedecke developed the study, collected data, and developed analyses. Kendra Hotz provided content review as a social scientist. Rachel Nelson, Hanah Walker, Anna Belle Gallaher, and Angela Nakahara provided content review as clinicians. Charisse Madlock and Lauren Camp provided structural and stylistic review. All authors reviewed the assembled document and suggested revisions, incorporated by Patricia Goedecke.

## Introduction

Historical redlining began as an attempt to buttress American financial stability shaken by the Great Depression. In 1933, nearly half of all American home mortgages were in default, and approximately one thousand homes per day were being foreclosed.^1^ President Roosevelt’s New Deal created the Home Owners’ Loan Corporation (HOLC) to provide mortgage refinance loans for borrowers struggling to make payments. To ensure the security of these investments for banks, the administration authorized HOLC to categorize neighborhoods by investment grade.^2^ A combination of local officials, real estate experts, and HOLC examiners assigned grades A through D to neighborhoods based on risk assessments.

Their maps were color-coded to reflect investment safety: The Grade A neighborhoods, considered the “best,” were green; Grade B areas were blue and “still desirable”; Grade C areas, “definitely declining,” were yellow; and the “hazardous” areas of Grade D were red. Red neighborhoods were often characterized by a larger proportion of minority residents.^3 4^ This scale came to be known as “redlining”—lending practices that systematically invested in predominantly White neighborhoods.^5 3 2^ While the HOLC ceased in 1951, unequal property investment and racialized perceptions of neighborhood value have persisted.

A growing body of research has investigated the relationship between HOLC neighborhood grades and current health outcomes.^6^ Studies show HOLC grades to be associated with poorer self-reported health, chronic conditions including hypertension and kidney disease, higher rates of 30-day post-operative mortality, and shorter life expectancy.^4 7 8 9 10^ While many researchers have modeled redlining as causal, others suggest it may be emblematic of a multifaceted history of structural racism that warrants further study. Some studies have examined mediating SDOH downstream of historic HOLC.^11^ Research suggests that the limited access to credit from historic redlining may have led to reduced home ownership and lower property values. This combination may have contributed to generational poverty, and with it, lack of educational achievement leading to poorer employment opportunities.^12 13 14^ Ultimately, these may have led to lack of access to resources, food insecurity, risk of eviction, and even heat vulnerability from lack of urban tree canopy.^15 16 17 18 19^

Hypertensive disorders of pregnancy (HDP) affect 2-8% of pregnancies in the United States (U.S.) and are a primary cause of maternal morbidities, including placental abruption, pulmonary edema, renal failure, and death. Black women in the U.S. face maternal mortality rates three times those of White women, from causes including HDP. In Memphis, Tennessee, where 64% of the population identifies as Black, HDP prevalence is notably high. Approximately 12% of patients who deliver at the city’s largest public tertiary hospital are diagnosed with HDP.^20 21^ Our first aim is to explore whether the prevalence of HDP in Memphis corresponds to historic HOLC grades.

The associations between social determinants and health outcomes are complex and rife with confounders, particularly where decades have elapsed between the two. Researchers have found significant relationships between HOLC grades and preterm delivery even when controlling for poverty levels and educational attainment, suggesting an impact beyond generational poverty.^15 22^ This prompts further investigation into how redlining has affected health: through economic impacts, physiological effects of chronic stress, or other means. We imagine some SDOH may have interwoven relationships with one another prior to impacting long-term health outcomes in the neighborhood; hence, our use of network modeling.

Recently released data shows current residential lending patterns still resemble those established by historic HOLC grades (**Figure 1**).^23^ Chronic stress is represented in community-level prevalence of chronic hypertension, an inflammatory illness shown to reflect allostatic load.^7 8 24^ HDP are associated with chronic hypertension both as sequelae and as risk factors; it is unclear whether HDP increases the risk of developing chronic hypertension or indicate pre-existing risk.^25^

**Figure 1.**
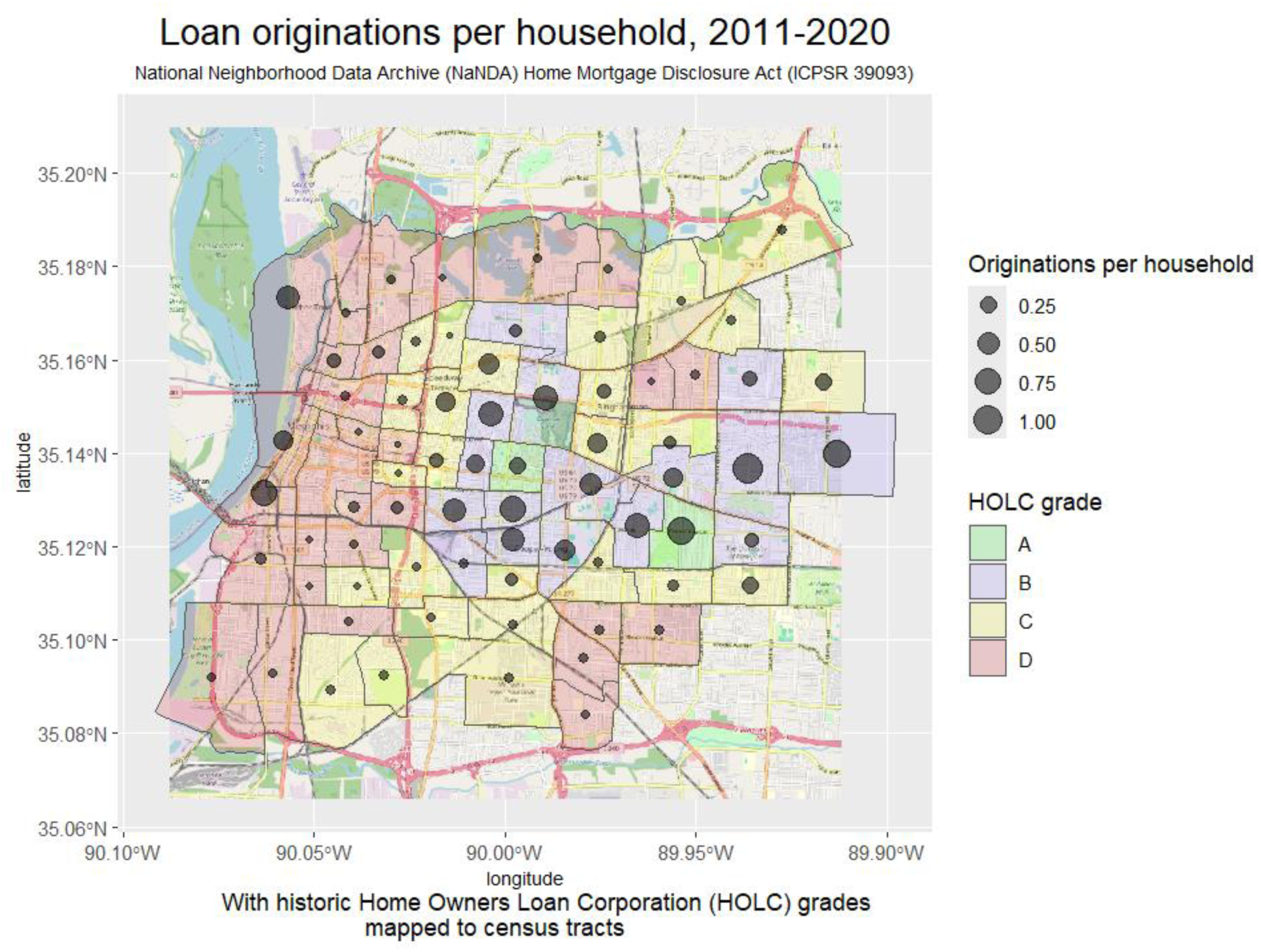
Map of HOLC grades with loan originations. Map of census tracts in Memphis with associated historical Home Owners’ Loan Corporation (HOLC) neighborhood grades and loan originations per household 2011-2020.

This study aims to validate existing research investigating relationships between historic redlining and contemporary obstetric outcomes; to assess whether the risk observed in other demographics also impacts mothers delivering in our city. We aim to explore which current SDOH are related to historic HOLC ratings, illuminating potential causal pathways. We hypothesize that (1) historic HOLC grades will demonstrate measurable relationships with HDP in Memphis, Tennessee; (2) HOLC grades will associate linearly with some of the current SDOH identified in literature as downstream effects of redlining; and (3) network modeling will elucidate more complex, interwoven relationships among SDOH which may eventually contribute to HDP. This network modeling is intended for exploratory use, to generate causal hypotheses which may stimulate further study rather than provide definitive answers regarding how historic Redlining Impacts Current Hdp.^26^

## Methods

### Study Population and Data Sources

We conducted a retrospective cohort study of singleton pregnancies delivered at one public and one private hospital system in Shelby County, Tennessee. Subjects were determined to have HDP if they received a diagnosis of gestational hypertension, preeclampsia, eclampsia, or hemolysis with elevated liver enzymes and low platelets (HELLP syndrome).^27^ We incorporated a spatially referenced map of historic HOLC grades with residential loan investment rates at census-tract level for 2011 through 2020.^28 23^ We included SDOH at the census-tract level, which was speculated in prior literature to be downstream effects of HOLC.^29 30^

Patients with singleton pregnancies were included in the study, who lived in the historically smaller Memphis city limits for which historic HOLC grades were available. Data from only the first pregnancy were retained for women with multiple pregnancies during the study date range. (Appendix 2).

### Objectives

Our first objective was to compare the prevalence of HDP by residence in historic neighborhoods grades A through D. Our second was to identify current SDOH, which may be downstream effects of redlining. Our third was to model network relationships among these current SDOH and HDP to explore the means by which redlining may impact current health.

### Study Design

This retrospective cohort study reviewed singleton deliveries among patients residing in neighborhoods historically graded A through D in Memphis, Tennessee, March 2019-December 2020. Deliveries took place within both a public and a private hospital system. The primary outcome was HDP, measured as a binary variable at the patient level and as a percentage of deliveries within each historical HOLC grade. Risk ratios for HDP were calculated per historic HOLC grade and compared using a chi-square test. Logistic associations with HDP at the patient level were measured first in univariate models for demographic, clinical, and social characteristics. The multivariable logistic model included variables with univariate significance and a Pearson correlation coefficient <0.60.

In a secondary analysis, we used prior literature to identify current SDOH which may represent downstream effects of redlining. We reassigned HOLC scores from A through D as numbers 1 through 4 and averaged the values within census tracts, with higher scores representing worse HOLC grades within an area. We developed univariate linear regression models with HOLC grades as predictors of current SDOH, in an attempt to establish the role of these current factors as downstream effects of redlining.

To explore complex interrelationships among current SDOH that impact HDP, we scaled our continuous variables and modeled these using Bayesian network analysis (BNA). BNA allows for the simultaneous modeling of relationships among a comprehensive set of variables, referred to as *nodes*. Probabilistic relationships between these nodes are represented by *arcs*. For the BNA algorithm, we applied the Max-Min Hill-Climbing (MMHC) algorithm within the R package *bnlearn*, which adapts well to small data sets and effectively models both categorical and continuous data. Initially, arcs are established and pruned using *mutual information* to distinguish between conditional dependence and conditional independence for each pair of nodes, conditioned on the full set of nodes. The strength of each arc indicates the degree to which one node influences another. Subsequently, parameters are estimated to represent the coefficients of linear relationships among continuous variables and conditional probabilities for categorical variables. Our BNA findings are visualized in a chord diagram, where the widths of the arcs indicate the strengths of the probabilistic relationships, with parameter estimates reflecting whether these relationships are positive or negative. We used an alpha of 0.05 as the criterion for inclusion of arcs in the graphical model and for all statistical significance. All analyses were performed using R 4.4.^31^

## Findings

We found 2371 patients residing in areas formerly classified to HOLC grades (). These mapped to 66 current census tracts, with a combined estimated 2020 population of 155,155. Demographic and clinical variables differed predictably by HOLC grade. Proportions of patients in each historic HOLC grade differed significantly in location of delivery (p<0.0001), with 17% of A-rated neighborhood residents, 27% of B-rated, 43% of C-rated, and 51% of D-rated neighborhood residents delivering at the public hospital (Table 1). Rates of HDP increased as HOLC grade level declined. Using grade D as a baseline, we found unadjusted risk ratios of .59, .55, and .77 (p=0.0129) for grades A through C, respectively. Preliminary logistic models of demographic, clinical, and SDOH risk factors for HDP found 6 were significant at the univariate level (Appendix 3). Of these, five were retained after excluding one for Pearson correlation >0.60. Our multivariable logistic model found an odds ratio of 1.22 (CI 1.00, 1.50), representing a 22% increase in HDP risk for each worsening HOLC grade level (Figure 2).

**Figure 2.**
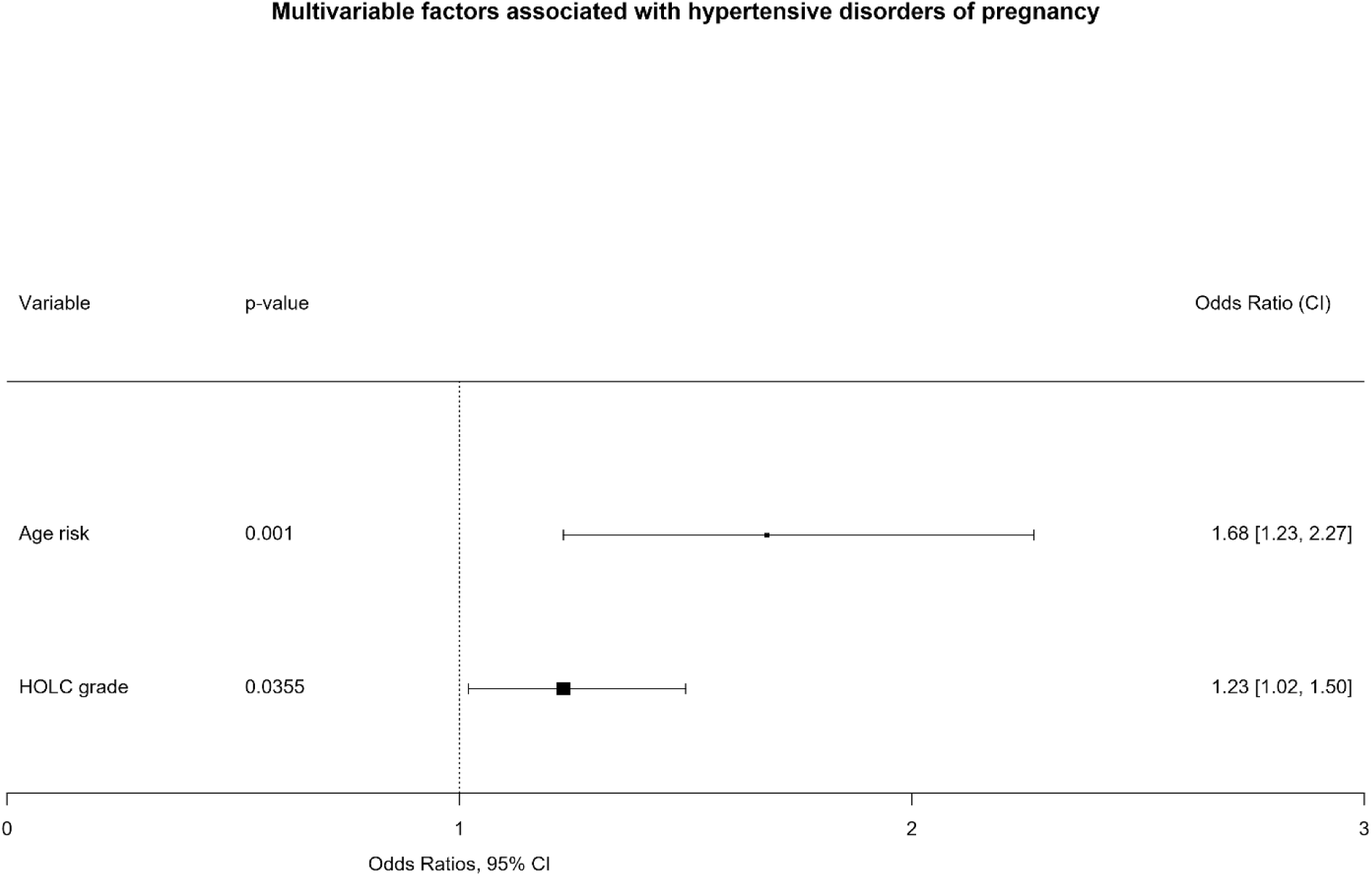
Forest plot. Forest plot showing odds ratios with 95% confidence intervals from stepwise multivariable logistic analysis associating demographic and clinical factors and historical Home Owners’ Loan Corporation (HOLC) neighborhood grades with patient-level hypertensive disorders of pregnancy (HDP).

**Table 1.** Demographic and clinical characteristics by HOLC grade. Demographic and clinical characteristics by historic Home Owners’ Loan Corporation (HOLC) grade. Shown are mean with standard deviation for continuous variables; count with percentage for categorical variables; p-values calculated using one-way analysis of means for continuous variables; Fisher’s exact test with categorical variables.

| Variable | A<br>N = 102 | B<br>N = 517 | C<br>N = 872 | D<br>N = 880 | p-value |
| --- | --- | --- | --- | --- | --- |
| Age |  |  |  |  | <0.001 |
| Mean (SD) | 32 (5) | 30 (6) | 27 (6) | 26 (6) |  |
| Age risk | 39 (38%) | 137 (26%) | 231 (26%) | 205 (23%) | 0.011 |
| Race |  |  |  |  |  |
| Black | 22 (22%) | 234 (45%) | 731 (84%) | 809 (92%) |  |
| White | 73 (72%) | 233 (45%) | 78 (8.9%) | 35 (4.0%) |  |
| Other | 7 (6.9%) | 50 (9.7%) | 63 (7.2%) | 36 (4.1%) |  |
| Race risk | 22 (22%) | 234 (45%) | 731 (84%) | 809 (92%) |  |
| HDP | 6 (5.9%) | 29 (5.6%) | 67 (7.7%) | 91 (10%) | 0.012 |
| Chronic Hypertension | 6 (5.9%) | 21 (4.1%) | 60 (6.9%) | 73 (8.3%) | 0.019 |
| Diabetes Type I or II | 0 (0%) | 1 (0.2%) | 1 (0.1%) | 4 (0.5%) | 0.6 |
| Drug Screen | 1 (1.0%) | 0 (0%) | 5 (0.6%) | 3 (0.3%) | 0.2 |
| IUFD | 1 (1.0%) | 2 (0.4%) | 7 (0.8%) | 9 (1.0%) | 0.5 |
| IUGR | 6 (5.9%) | 8 (1.5%) | 7 (0.8%) | 7 (0.8%) | 0.002 |
| Kidney Disease | 1 (1.0%) | 4 (0.8%) | 1 (0.1%) | 2 (0.2%) | 0.10 |
| Smoker | 3 (2.9%) | 53 (10%) | 152 (17%) | 129 (15%) | <0.001 |
| Thrombophilia | 3 (2.9%) | 2 (0.4%) | 2 (0.2%) | 0 (0%) | <0.001 |
| Preterm Delivery | 7 (6.9%) | 29 (5.6%) | 69 (7.9%) | 93 (11%) | 0.011 |
| Hospital |  |  |  |  | <0.001 |
| Private hospital | 85 (83%) | 372 (72%) | 496 (57%) | 422 (48%) |  |
| Public hospital | 17 (17%) | 145 (28%) | 376 (43%) | 458 (52%) |  |

For our second aim, we used prior literature to identify 8 current SDOH as potential downstream effects of redlining. Although we did not anticipate strictly linear relationships between HOLC grades and these effects, we found 5 of the 7 continuous SDOH to have linear relationships with HOLC grades (Table 2). Our BNA analysis did not show significant network relationships between current SDOH and patient-level HDP.

**Table 2.** Univariate linear associations with HOLC ratings. Univariate linear associations of SDOH and chronic hypertension with historic Home Owners’ Loan Corporation (HOLC) ratings averaged within census tracts

| **Variable** | **Coefficient** | **SE** | **p-value** |
| --- | --- | --- | --- |
| Chronic hypertension | 6.03 | 1.29 | <0.001 |
| Housing loan percent | -19.67 | 3.08 | <0.001 |
| Home improvement percent | -0.62 | 0.12 | <0.001 |
| Service industry | 4.37 | 1.30 | 0.0013 |
| Vacancy rate | 4.68 | 1.19 | <0.001 |
| Neighborhood growth | -2.95 | 2.67 | 0.2747 |
| Deep poverty | 5.71 | 1.59 | <0.001 |
| Loan leverage ratio | -0.07 | 0.14 | 0.6113 |

a Averaged HOLC grade predicting linear outcomes; A=1, B=2, C=3, D=4

Our BNA did suggest linear relationships from scarcity of residential loans to population-level chronic hypertension, to property abandonment. Other arcs within the model suggested relationships from property abandonment to deep poverty and lack of neighborhood growth; from food deserts to reduced loan leverage ratios, to diminished neighborhood growth; and from scarcity of residential loans to employment in the service industry. In the chord diagram, the widths of arcs indicate probabilistic strengths of relationships; linear coefficient estimates indicate positive or negative direction and unit scale (Figure 3, Table 3). We consider this BNA to be exploratory rather than definitive in nature, intended to stimulate further exploration and discussion.

**Figure 3.**
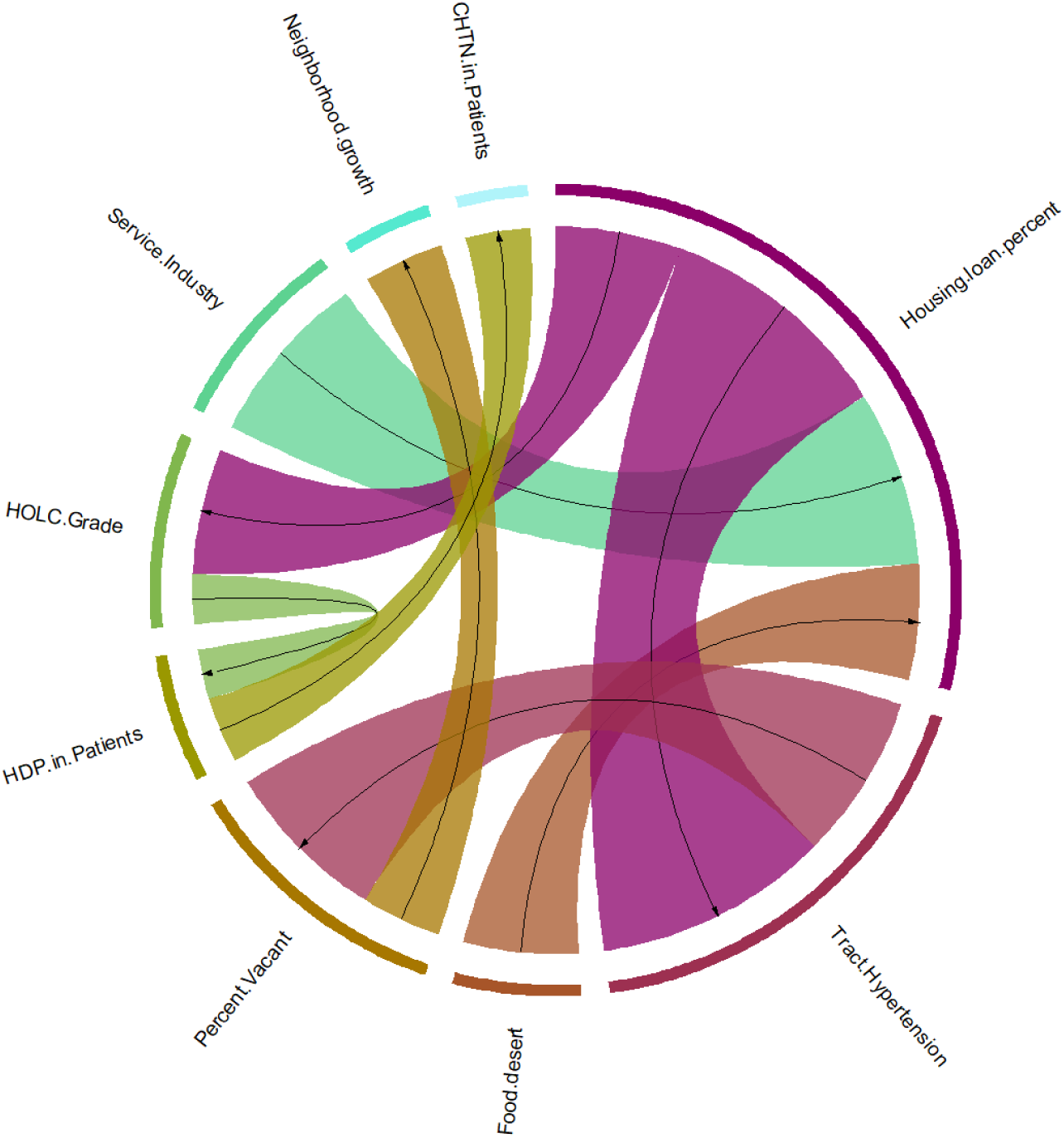
Graphical model of Bayesian Network Analysis. Graphical model of Bayesian Network Analysis using the Max-Min Hill Climbing algorithm to examine strengths of probabilistic relationships among historical Home Owners’ Loan Corporation (HOLC) neighborhood grades, current factors at the census-tract level, and the prevalence of hypertensive disorders of pregnancy (HDP) and chronic hypertension (CHTN) among women delivering 2019-2020 at a public and private hospital in Memphis, TN. Arc strengths and parameter estimates are shown in Table 3.

**Table 3.** Bayesian network analysis. Arc strengths and parameter estimates from Bayesian network analysis of census-tract level social determinants of health (SDOH) related to historic Home Owners’ Loan Corporation (HOLC) ratings. Network determined using Max-Min Hill-Climbing (MMHC) local network learning algorithm for hybrid discrete and continuous data, at alpha=0.05.

| From Node | To Node | Strength | Estimate |
| --- | --- | --- | --- |
| Housing loan percent | Tract Hypertension | 0.47 | -0.78 |
| Service Industry | Housing loan percent | 0.35 | -0.16 |
| Tract Hypertension | Percent Vacant | 0.33 | 0.70 |
| Housing loan percent | HOLC Grade | 0.25 | -0.62 |
| Food desert | Housing loan percent | 0.23 | -0.73 |
| Percent Vacant | Neighborhood growth | 0.16 | -0.52 |
| HDP in Patients | CHTN in Patients | 0.13 | 0.47 |
| HOLC Grade | HDP in Patients | 0.10 | 0.43 |

## Discussion

Our primary analysis shows a clear relationship between historic HOLC grades and current health outcomes as found in prior studies. What remains less clear is how this relationship is mediated. Our network analysis suggests that continued disinvestment, as evidenced by scarce residential loan availability and vacant properties, may be related to the prevalence of chronic hypertension. We consider this model to be exploratory in nature, generating hypotheses which may be worthy of further exploration.

The linkage between historic HOLC grades and current health outcomes remains largely unexplained. As instructed at the time, banks reduced investment in poorly graded communities. Analyses have often modeled redlining as directly affecting health outcomes; some researchers have posited potential causal pathways such as racial segregation, capital disinvestment, or reduced wealth accumulation from homeownership. Others suggest that the HOLC system did not cause systemic problems but simply reflected existing social realities—perhaps cementing realities of that time, or perhaps color-coding realities that are just as much of our time.^11 32^

Our study validates the ongoing obstetric health risk of living in neighborhoods historically graded poorly by HOLC. Our measurement of linear relationships between HOLC grades and current SDOH lends credibility to the theory of these as downstream effects of HOLC which may act as mediators impacting health today. We failed to identify a pathway by which HOLC impacts HDP via current SDOH. The BNA relationships found for population-level chronic hypertension with current scarcity of residential lending and abundance of property vacancies may be interpreted as lending validity to economic and/or psychological factors as mediators between HOLC and health.

This study is limited by its small geographic scope to a single metropolitan area and its focus on a health outcome that impacts a small portion of the population at any given time (pregnant individuals). Additionally, BNA of potential downstream effects of a decades-old government initiative does not lend itself to definitive answers; rather, it is a method of exploration intended to stimulate further analysis.

Strengths of our study include taking a step beyond the assumption that HOLC grades could directly impact current health outcomes, as well as not limiting our expectations to any one mediating effect of the HOLC grading system. We focused on a metropolitan area with a high prevalence of HDP and chronic hypertension, where the historical impacts of racial segregation are well documented.

Our findings validate prior studies showing that historical HOLC grades impact current hypertensive disorders, particularly during pregnancy. We recommend that further research is necessary to understand how exactly historic HOLC grades still impact health today. Improved understanding of the intervening factors may lead to strategic investments in neighborhoods historically affected by poor HOLC ratings, to mitigate downstream effects of this rating system on current health.

## Data Availability

All data produced in the present work are contained in the manuscript.

---

**Appendix 2.** Inclusions / Exclusions.

| Private hospital |  | Public hospital |  | REASON |
| --- | --- | --- | --- | --- |
| INCLUDE | EXCLUDE | INCLUDE | EXCLUDE |  |
| 115,096 |  | 4758 |  | Original data |
| 33,078 | 81,994 | 4758 | - | Include only singleton deliveries<br>2019 March through 2020 December |
| 6429 | 26,649 | 3508 | 1250 | Exclude non-residential properties,<br>outside Shelby County, missing<br>street address or failed to merge to<br>latitude/longitude |
| 1375 | 5054 | 996 | 2512 | Exclude those outside the historical<br>boundaries of Memphis rated using<br>the Home Owners' Loan Corporation<br>(HOLC) grade scale |

**Appendix 3.**
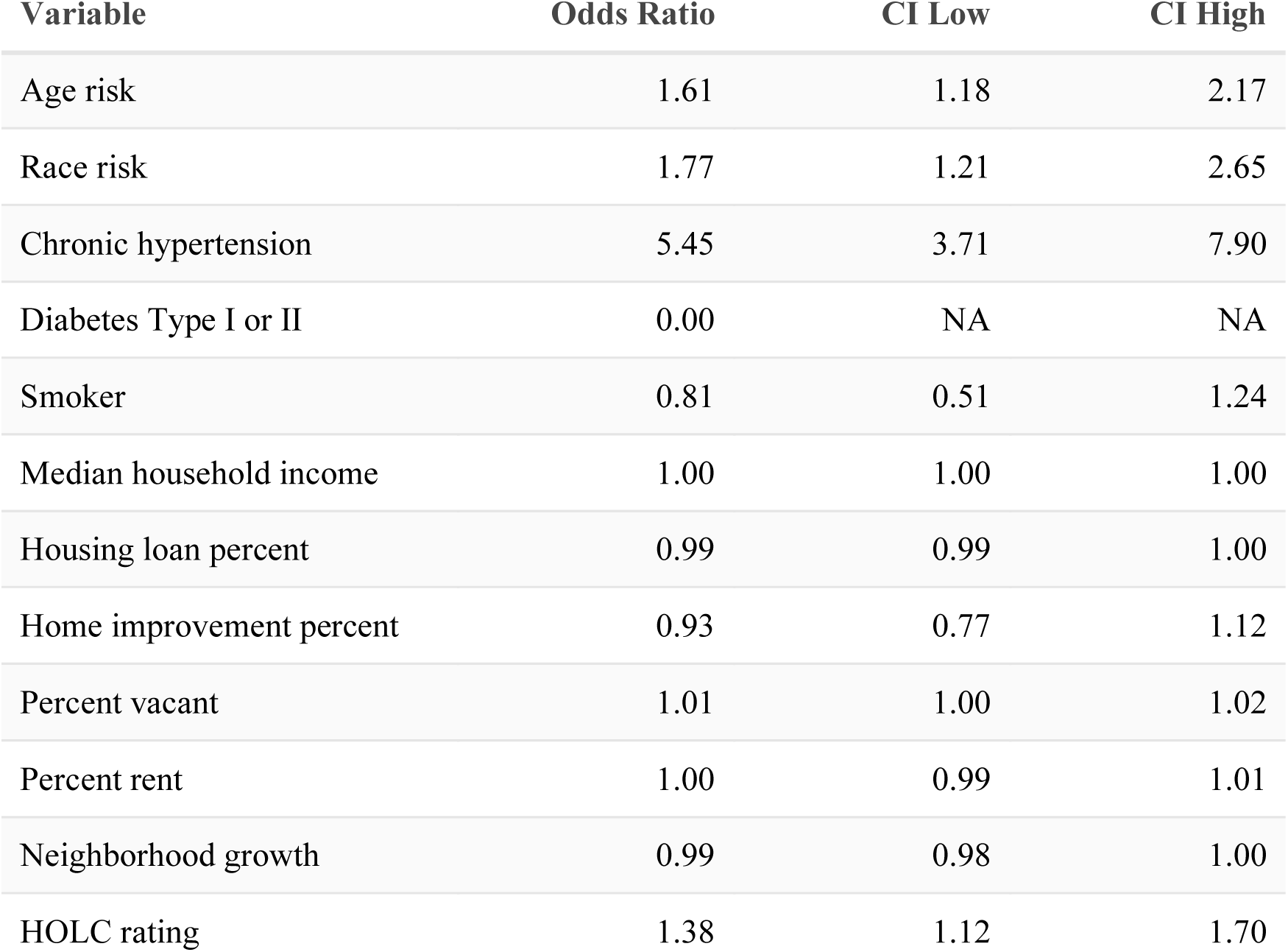
Univariate logistic associations with HDP. Univariate logistic associations with hypertensive disorders of pregnancy (HDP) at patient level, Memphis, Tennessee, 2019-2021

| Variable | Odds Ratio | CI Low | CI High |
| --- | --- | --- | --- |
| Age risk | 1.61 | 1.18 | 2.17 |
| Race risk | 1.77 | 1.21 | 2.65 |
| Chronic hypertension | 5.45 | 3.71 | 7.90 |
| Diabetes Type I or II | 0.00 | NA | NA |
| Smoker | 0.81 | 0.51 | 1.24 |
| Median household income | 1.00 | 1.00 | 1.00 |
| Housing loan percent | 0.99 | 0.99 | 1.00 |
| Home improvement percent | 0.93 | 0.77 | 1.12 |
| Percent vacant | 1.01 | 1.00 | 1.02 |
| Percent rent | 1.00 | 0.99 | 1.01 |
| Neighborhood growth | 0.99 | 0.98 | 1.00 |
| HOLC rating | 1.38 | 1.12 | 1.70 |

**Appendix 4.** Demographic and clinical characteristics by hospital. Shown are mean with standard deviation for continuous variables, counts with percentage for categorical variables.

| Characteristic | Private<br>N = 1375 | Public<br>N = 996 | p-value |
| --- | --- | --- | --- |
| Age |  |  | <0.001 |
| Mean (SD) | 28 (6) | 26 (6) |  |
| Age risk | 382 (28%) | 230 (23%) | 0.010 |
| Race |  |  | <0.001 |
| Black | 943 (69%) | 853 (86%) |  |
| White | 333 (24%) | 86 (8.6%) |  |
| Other | 99 (7.2%) | 57 (5.7%) |  |
| Race risk | 943 (69%) | 853 (86%) | <0.001 |
| Chronic Hypertension | 59 (4.3%) | 101 (10.0%) | <0.001 |
| Diabetes Type I or II | 6 (0.4%) | 0 (0%) | <0.001 |
| Drug Screen | 2 (0.1%) | 7 (0.7%) | 0.040 |
| IUFD | 17 (1.2%) | 2 (0.2%) | 0.005 |
| IUGR | 26 (1.9%) | 2 (0.2%) | <0.001 |
| Kidney Disease | 5 (0.3%) | 3 (0.3%) | >0.9 |
| Smoker | 315 (23%) | 22 (2.2%) | <0.001 |
| Thrombophilia | 6 (0.4%) | 1 (0.1%) | 0.3 |
| Preterm Delivery | 5 (0.4%) | 193 (19%) | <0.001 |
| HDP | 72 (5.0%) | 129 (12%) | <0.001 |
| HOLC grade |  |  | <0.001 |
| A | 85 (6.2%) | 17 (1.7%) |  |
| B | 372 (27%) | 145 (15%) |  |
| C | 496 (36%) | 376 (38%) |  |
| D | 422 (31%) | 458 (46%) |  |

