## Appendix 1 for "Historic Redlining and Hypertensive Disorders of Pregnancy: A Retrospective Cohort Study"

Appendix 1: Data dictionary

| HDP[1] Risk factor | Category | Source level | Definition/Source |
|---|---|---|---|
| Age risk (age <17 or age>35) | Clinical | EHR[2] individual | |
| Chronic Hypertension | Clinical | EHR individual | |
| Diabetes Type I or II | Clinical | EHR individual | |
| Kidney Disease | Clinical | EHR individual | |
| Smoker | Clinical | EHR individual | |
| Race risk ie Black | Clinical | EHR individual | |
| Hospital | SDOH | EHR individual | |
| Deep Poverty | SDOH | Census tract | Estimated percent of families that live in deep poverty (at less than 50% of the poverty level), between 2014-2018. PolicyMap. Based on data from Census: US Bureau of the Census, American Community Survey. Accessed 15 January 2020. http://www.policymap.com.\ |
| Home improvement percent | SDOH | Census tract | Percent of households receiving loan originations for home improvement purposes, 2011-2020. Edlebi, Jad, Mitchell, Bruce, Richardson, Jason, Meier, Helen, Chen, Liang, Noppert, Grace, and Gypin, Lindsay. National Neighborhood Data Archive (NaNDA): Home Mortgage Disclosure Act Longitudinal Dataset by Census Tract, United States, 1981-2021. Inter-university Consortium for Political and Social Research [distributor], 2024-05-15. https://doi.org/10.3886/ICPSR39093.v1 |
| Housing loan percent | SDOH | Census tract | Percent of households receiving loan originations for residential purposes, 2011-2020. Edlebi, Jad, Mitchell, Bruce, Richardson, Jason, Meier, Helen, Chen, Liang, Noppert, Grace, and Gypin, Lindsay. National Neighborhood Data Archive (NaNDA): Home Mortgage Disclosure Act Longitudinal Dataset by Census Tract, United States, 1981-2021. Inter-university Consortium for Political and Social Research [distributor], 2024-05-15. https://doi.org/10.3886/ICPSR39093.v1 |
| Hypertension | SDOH | Census tract | Crude percent of high blood pressure among adults aged >=18 years in 2017. PolicyMap. Based on data from CDC_PLACES: Data downloaded from https://www.cdc.gov/places/index.html. Accessed 15 January 2020. http://www.policymap.com. |
| Loan Leverage Ratio | SDOH | Census tract | Median loan to income ratio. PolicyMap, Federal Financial Institutions Examination Council (FFIEC): Home Mortgage Disclosure Act (HMDA) Summaries. Updated October 2023. http://www.ffiec.gov/hmda/ |
| Neighborhood growth | SDOH | Census tract | Percent change in the number of housing units occupied from 2010 to 2020. US Census table S2504. |
| Never Married | SDOH | Census tract | Estimated percent of people age 15 and over who have never been married, between 2014-2018. PolicyMap. Based on data from Census: US Bureau of the Census, 2000 Longform. Accessed 15 January 2020. http://www.policymap.com. |
| Percent Rent | SDOH | Census tract | Estimated percent of renter-occupied housing units, between 2014-2018. PolicyMap. Based on data from Census: US Bureau of the Census, American Community Survey. Accessed 15 January 2020. http://www.policymap.com. |
| Service Industry | SDOH | Census tract | Estimated percent of people age 16 years or older who were employed in the Accommodation and Food Services industry, between 2014-2018. PolicyMap. Based on data from Census: US Bureau of the Census, American Community Survey. Accessed 15 January 2020. http://www.policymap.com. |
| Vacancy Rate | SDOH | Census tract | Estimated percent of all housing units that were vacant type "other" in 2014-2018. PolicyMap. Based on data from Census: US Bureau of the Census, American Community Survey. Accessed 15 January 2020. http://www.policymap.com. |

NOTE: EHR variables included are those available in both public and private hospital records

[1] HDP: Hypertensive Disorders of Pregnancy

[2] EHR: Electronic Health Record
